# Trajectories and characteristics of pain in the first year after spinal cord injury: a TRACK-SCI study

**DOI:** 10.64898/2026.10.21.26366160

**Authors:** Jason H. Gumbel, Esmeralda Mendoza, Mayra Arellano, J. Russell Huie, Debra D. Hemmerle, Lisa U. Pascual, Vineeta Singh, Anastasia V. Keller, Xuan Bradfield, Philip R. Weinstein, Jacqueline C. Bresnahan, Michael S. Beattie, Adam R. Ferguson, John L.K. Kramer, Nikos Kyritsis, the TRACK-SCI Investigators

## Abstract

Chronic pain affects most individuals with spinal cord injury (SCI) and frequently impairs daily function, and nearly half experience neuropathic pain. Neuropathic pain is commonly thought to develop months after injury and to persist once established. These views rest on classifying each patient as having neuropathic pain or not, although patients often report several pains. The same binary label is the usual target of prognostic and biomarker studies that aim to predict life-limiting pain. We tracked up to three of the worst pains per interview from injury to one year in 140 TRACK-SCI participants, using the DN4 interview (DN4i, threshold 3). Within three days of injury, half of all pain areas and three quarters of participants met the threshold, and pain-area prevalence changed little over the year. Individual pains, however, often changed. Only a third remained above the threshold throughout, one third resolved or fluctuated, and pains in the same person often followed different courses. At more than half of interviews with several pains, one pain met the threshold while another did not, so that the patient’s classification depended on the pain considered. We therefore derived a threshold-free Pain Composite Score from the DN4i items and pain intensity. The items formed a single dimension, and the composite was associated with sleep and mood interference whereas the binary classification was not. Neuropathic pain after SCI thus appears early, does not invariably persist, and differs between a person’s pains, and graded pain-level measures may offer a more stable target for predicting life-limiting pain.

**Summary:** Neuropathic pain after spinal cord injury appears early and does not always persist. A threshold-free score, unlike binary classification, tracks how pain limits daily life.

## 1. Introduction

Chronic pain affects up to 80% of individuals with spinal cord injury (SCI) and reduces their quality of life [14,37,38], and about half of all individuals with SCI develop neuropathic pain [9,36]. Neuropathic pain arises from a lesion or disease of the somatosensory nervous system [41]. The International Spinal Cord Injury Pain (ISCIP) classification divides it into at-level and below-level pain relative to the neurological level of injury (NLI), a distinction thought to reflect different mechanisms [8,15,37,38].

Current understanding of neuropathic pain after SCI rests on two assumptions. The first is that it develops over the months following injury, which is why it is usually assessed in the subacute and chronic phases, even though it can be detected within days [34] and is preceded by altered pain modulation in the first weeks [12,19]. The second is that neuropathic pain persists once it is established, a view supported by its persistence between one and six months in pooled cohorts [44] and by its stable prevalence during inpatient rehabilitation [10]. Both assumptions derive from dichotomous classifications of the patient or of the single most bothersome pain, the pain that clinical studies usually assess [46], and group estimates built on such classifications may conceal change in individual pains. Pain intensity follows distinct trajectories during rehabilitation [7], and individuals often report several pains that may each follow their own course, so that the pain a patient ranks as worst may not be the one with the most neuropathic features.

How neuropathic pain is measured may in turn shape what is concluded about its course. In research settings, it is usually identified with screening questionnaires such as the Douleur Neuropathique en 4 Questions (DN4), whose seven-item interview version (DN4i) retains acceptable accuracy at a threshold of 3 when the examination items cannot be obtained [6], and which reached an area under the curve of 0.86 in SCI [20]. Such questionnaires were designed to support rather than replace a clinical diagnosis based on the description, location, and examination of pain [8,16,29,31], and were validated almost exclusively at their cutoff [6,13,30,31], where specificity can be modest [13]. Used as binary classifications, they place pains that differ by a single item on opposite sides of the decision, so that small fluctuations appear as new or resolved pain and statistical power is lost in prognostic and biomarker studies [1]. A continuous score would avoid these problems, but whether the DN4 items reflect a single dimension, the property that justifies summing them, has rarely been examined [39], and instruments that grade symptom severity [5,49] are seldom collected in acute cohorts. Moreover, neither a binary classification nor a graded symptom score indicates whether a pain limits daily life, which is what matters most to patients.

These limitations matter most for studies that aim to predict neuropathic pain. Prognostic models and biomarker studies require an outcome that is defined consistently across patients and over time, and they have generally labeled patients as developing or not developing neuropathic pain, on the assumption that the condition persists once present [45]. When patients report several pains with different characteristics, and when a change in a single item, such as a newly reported tingling, can move a pain across the threshold between interviews, such a label may depend on which pain is assessed and when. A model trained to recognize it may then learn the instability of the classification rather than differences in the underlying pain. Prediction may therefore be better served by a graded target that reflects both the neuropathic character of a pain and its intensity, the features most likely to determine whether a pain limits a patient’s life.

To our knowledge, no study has followed each pain separately from injury. We therefore tracked each reported pain, up to three per interview, in the Transforming Research and Clinical Knowledge in SCI (TRACK-SCI) study from the hyperacute phase to one year, to ask whether early onset and persistence hold for individual pains rather than for patients. We then derived a threshold-free Pain Composite Score from the DN4i items and pain intensity and examined whether the items form a single dimension. Finally, we asked whether the pain that participants ranked as worst was the pain with the most neuropathic characteristics, and related the composite score, the binary classification, and pain intensity to interference with sleep, mood, and day-to-day activities.

## 2. Methods

### 2.1. Study design and participants

The data were collected in TRACK-SCI, an ongoing multicenter prospective observational study conducted by the Brain and Spinal Injury Center (BASIC) at the University of California San Francisco (ClinicalTrials.gov NCT04565366) [42]. The study was approved by the institutional review board of the University of California San Francisco (protocol 15-16115, Transforming Research and Clinical Knowledge in Spinal Cord Injury), and participants or their legally authorized representatives provided informed consent. Only participants enrolled at Zuckerberg San Francisco General Hospital and Trauma Center (UCSF/ZSFG) routinely received in-hospital pain assessments, and the present analysis was therefore restricted to that site. Participants were assessed with the DN4 during the acute hospital stay, on the same days as the International Standards for Neurological Classification of SCI (ISNCSCI) examinations (days 0 to 7 and discharge) [35,42], and at 3, 6, and 12 months after injury by structured interview. A comprehensive list of TRACK-SCI measures has been published previously [42]. Age at injury, gender, race, ethnicity, and mechanism of injury were taken from the TRACK-SCI database, and the American Spinal Injury Association Impairment Scale (AIS) grade and the NLI were taken from the ISNCSCI examinations. Table 1 reports the AIS grade and the level of injury at the last in-hospital ISNCSCI examination. Participants were included if they had at least one DN4 assessment in which all seven interview items were answered, irrespective of the examination items.

**Table 1.**
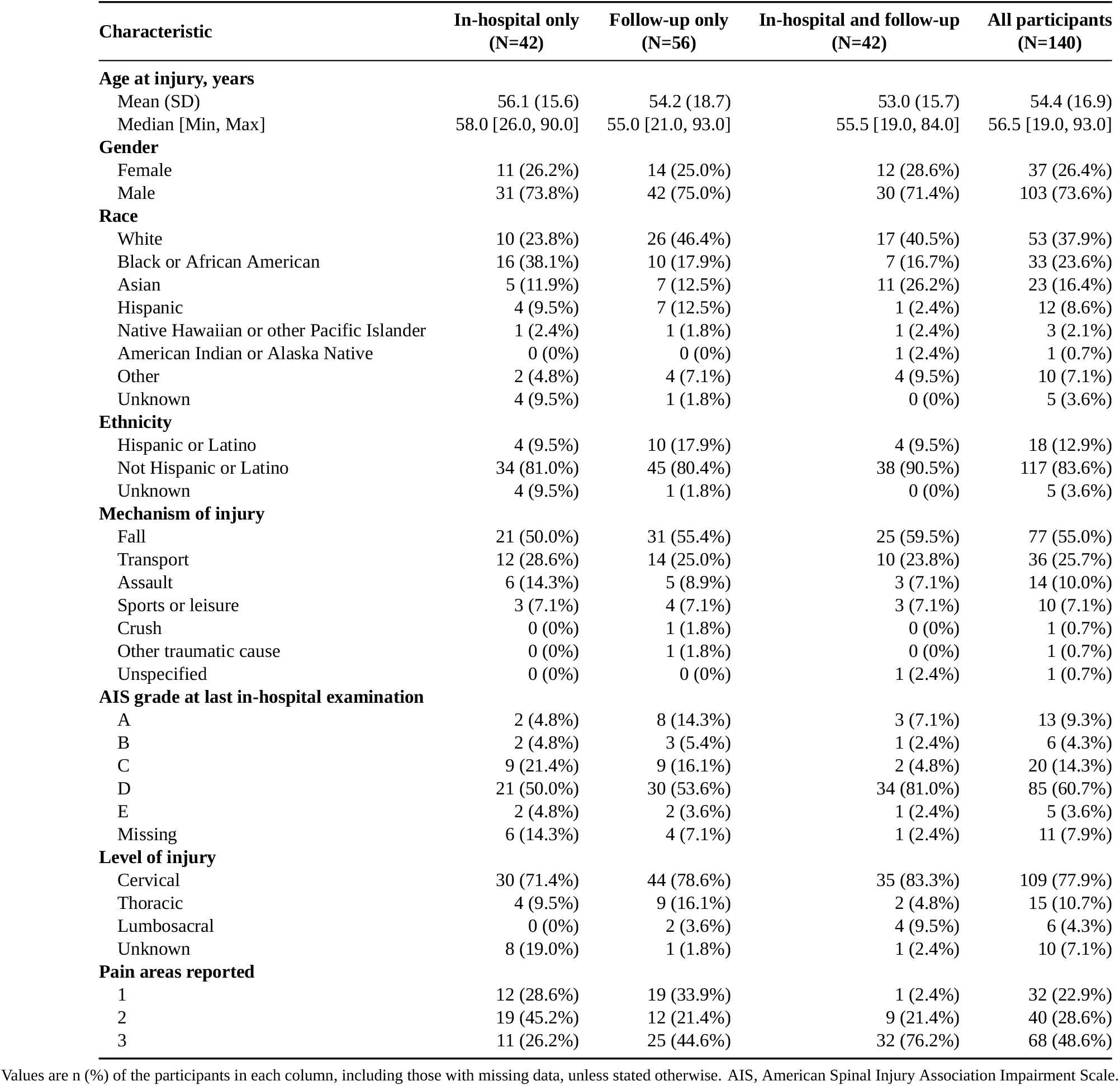
Cohort characteristics by the assessment phases each participant contributed.

### 2.2. Pain assessment

Pain was assessed with a modified version of the International SCI Pain Basic Data Set [46] in structured interviews, conducted in person during the acute hospitalization and almost always by telephone at follow-up, apart from occasional interviews conducted in person at a follow-up ISNCSCI examination. At each interview, participants described up to three pain problems, hereafter called pain areas, numbered 1 to 3, and ranked them from worst to least severe. The number followed the pain across interviews (Section 2.4), independently of its rank. For each pain area, they rated the average pain intensity over the previous week on a 0 to 10 numeric rating scale and completed the DN4. At each follow-up interview, participants also rated how much their pain had interfered with day-to-day activities, overall mood, and sleep in the previous week, each on a 0 to 10 scale. Current pain treatments and their drug classes were recorded at follow-up.

For pain area 1 at the 6- and 12-month interviews, pain location relative to the NLI was classified as above, at, or below level by applying the ISCIP definition [8] to the body regions recorded on the pain location checklist. The reference level was the NLI from the ISNCSCI examination at the same time point, or the estimated NLI recorded when the examination could not be performed (one interview). The at-level window comprised the NLI dermatome and the three dermatomes below it. Each body region was mapped to its approximate dermatome range, and a pain area was classified as below-level when any region lay more than three dermatomes below the NLI, even if the pain also involved the at-level zone, as at-level when its regions fell within the window, and as above-level when all regions lay rostral to the NLI. Pain areas with a region straddling the lower edge of the window could not be placed unambiguously and were classified as at-level.

### 2.3. DN4 interview scoring and prevalence

The DN4 comprises seven symptom descriptors, such as burning, tingling, and numbness, and three items that require a physical examination [6]. Because most assessments were performed by telephone, the examination items could not be obtained reliably, and all analyses used the seven-item interview version (DN4i). In patients after fracture surgery, a DN4i administered by telephone gave the same classification as an earlier in-person DN4 in 94% of cases [43]. Each item was scored 1 (yes) or 0 (no), giving a total of 0 to 7, and a score of 3 or more was considered screen-positive [6]. A pain area with a screen-positive score is referred to as DN4i-NP, a term that denotes the questionnaire classification rather than a clinical diagnosis of neuropathic pain.

Prevalence of DN4i-NP was calculated for pain areas and for participants (any pain area positive) in five time bins (0 to 3 days, 4 to 14 days, 3, 6, and 12 months). Because some pain areas were assessed more than once during the hospital stay, each pain area was counted once per time bin and considered positive when any of its assessments in that bin were positive. The 95% confidence intervals were exact binomial intervals for participants and, to account for the nesting of pain areas within participants, bootstrap intervals that resampled participants (5,000 resamples) for pain areas. At interviews with more than one pain area, we also recorded whether all, some, or none of the pain areas were DN4i-NP, and the participant-level classification (any pain area positive) was followed across time bins in participants assessed in at least two bins.

### 2.4. Pain-area trajectories

Trajectories were defined for individual pain areas. Because the order in which participants reported their pains could change between interviews, each pain area was matched across interviews by its reported location, so that a trajectory followed the same pain over time. For each pain area with at least one in-hospital assessment (0 to 14 days) and at least one follow-up assessment, we constructed a status sequence across the five time bins used for prevalence, in which a bin was screen-positive when any assessment within it was screen-positive. A sequence with no change in status was classified as Persistent DN4i-NP (all positive) or No DN4i-NP (all negative), a single change as Resolved DN4i-NP (positive to negative) or Delayed-onset DN4i-NP (negative to positive), and two or more changes as Fluctuating DN4i-NP. Trajectories were defined by these rules rather than by data-driven clustering because the number of pain areas and of assessments per pain area was too small to estimate latent trajectory classes reliably. Because pain areas assessed more often had more opportunities to change, we also report how often the status changed from one assessed time bin to the next, a measure that does not depend on the number of assessments. In a sensitivity analysis, the status of each in-hospital bin was taken from its last assessment. A second analysis classified all pain areas with at least two follow-up assessments, using the same rules without the in-hospital status.

### 2.5. Pain Composite Score and dimensionality of the DN4i

Because the DN4 was designed as a binary screening tool and a binary classification is poorly suited to prognostic and biomarker models, we derived a threshold-free score. Pain intensity was included alongside the seven items so that the score would reflect how severe each pain was as well as its neuropathic character. The seven DN4i items and average pain intensity were standardized and entered into a principal component analysis (PCA), following the syndromic workflow previously described for neurotrauma data and implemented in the syndRomics package [40]. We used linear rather than nonlinear PCA because a binary item admits only one quantification, so that nonlinear PCA yields the same solution for the seven items [28], whereas its rescaling of pain intensity would depend on the number of components extracted. The PCA was fitted on the 528 pain-area assessments with a recorded intensity, and scores were computed for all 596 assessments by setting a missing intensity to the sample mean. This applied to 68 assessments, most of them at follow-up interviews that also lacked interference ratings, and to only 2 assessments in the interference models. The number of components to retain was determined with the permutation test implemented in syndRomics [40]. In an approach analogous to Horn’s parallel analysis [22], this test compares the variance accounted for (VAF) by each component with its distribution after 10,000 permutations of the data, with Benjamini-Hochberg-adjusted p-values across all components [3]. Loadings were standardized for interpretation. The first component (PC1) was oriented so that DN4i items loaded positively, scaled to unit standard deviation, and retained as the Pain Composite Score. The relationship between PC1, the DN4i total, and pain intensity was described with Spearman correlations.

Because combining the DN4i items into a single score assumes that they reflect one underlying dimension, the PCA and permutation test were repeated with the seven DN4i items alone, using all pain-area assessments with complete items. To address the nesting of pain areas within participants, this analysis was also performed with one pain area per participant (the first assessment of the worst-ranked pain). The internal consistency of the summed items was then estimated with the Kuder-Richardson formula 20 (KR-20).

### 2.6. Worst-ranked pain and neuropathic characteristics

At interviews in which more than one pain area was reported, we calculated the proportion of interviews at which the participant’s worst-ranked pain had the highest PC1, DN4i total, and pain intensity among that participant’s pain areas, counting ties as concordant. The expected proportion under random ranking was the mean of 1/k across interviews, where k is the number of pain areas.

### 2.7. Association with pain interference

Associations with pain interference were tested at the 3-, 6-, and 12-month interviews for the pain that participants themselves ranked as worst. Because interference was rated for pain overall, this is the pain most likely to limit daily life, and it is identified independently of its neuropathic characteristics. Selecting the pain with the highest PC1 would have made the analysis circular, and averaging across pain areas would have diluted the contribution of the pain that limits the patient most. For each interference domain (sleep, mood, day-to-day activities), a linear mixed model was fitted with PC1 as the outcome, with interference, time point, and their interaction as fixed effects, any current pain treatment and the AIS grade at the time point as covariates, and a random intercept per participant [2]. The interference effect was tested with a Type III F-test. With the interaction in the model, this test evaluates the interference slope averaged across the three time points, and it does not depend on the coding of the time factor. Averaged and time point-specific slopes with 95% confidence intervals were estimated with estimated marginal trends [27], and the time point-specific slopes are presented descriptively. To display the data underlying each model, predicted PC1 values across the interference range were estimated at each time point, averaging over the covariates in proportion to their frequency. P-values for the three domains were adjusted for multiple testing as described in Section 2.9.

Four further analyses characterized these associations. First, a model without the interaction served as a sensitivity analysis. Second, each participant’s interference score was decomposed into the participant mean (between-participant component) and the deviation from it (within-participant component), which were entered together [11]. Third, to compare scoring approaches, the models were refitted in a common sample with standardized outcomes and standardized interference, using in turn PC1, the DN4i total, the binary DN4i classification (score of 3 or more), and pain intensity as the outcome. Finally, interference was modeled as the outcome with standardized intensity and PC1 entered together, with and without the number of pain areas. Because PC1 incorporates pain intensity, this model was repeated with the DN4i total, which does not, in place of PC1.

### 2.8. Opioid use

Opioid use at each follow-up interview was classified as present when an opioid was recorded for any pain area and absent only when the medication record documented no opioid use. Interviews at which the medication list could not be obtained were treated as unknown and excluded. Because few treated participants had confirmed absence of opioid use, this comparison is reported descriptively (Fig. S6B).

### 2.9. Statistical analysis

Continuous variables were summarized as mean and standard deviation or median and range, and categorical variables as counts and percentages. Prevalence, trajectories, the concordance between the worst-ranked pain and its neuropathic characteristics, and the analyses of pain location and opioid use were descriptive. Inferential analyses comprised the permutation tests of the principal components (Section 2.5) and the linear mixed models of pain interference (Section 2.7). Mixed models were fitted by restricted maximum likelihood, and fixed effects were tested with Type III F-tests using Satterthwaite degrees of freedom [25]. Because participants contributed several pain areas and several time points, this dependence was accounted for by the participant random intercept in the mixed models and by resampling participants rather than assessments for the bootstrap confidence intervals. Each model used all assessments with complete data for its variables, and the only imputation was the setting of a missing pain intensity to the sample mean when PC1 was computed (Section 2.5). Model residuals and random effects were inspected for normality and constant variance, and the influence of individual participants was assessed by refitting each model with one participant omitted at a time. Benjamini-Hochberg adjustment [3] was applied across components in the permutation tests and across the three interference domains in the primary mixed models, whereas the sensitivity and secondary analyses were not adjusted for multiple testing. Two-sided p-values below 0.05 were considered statistically significant. Analyses were performed in R 4.6.0 with the packages syndRomics 0.1.0 [40], lme4 2.0-6 [2], lmerTest 3.2-1 [25], emmeans 2.0.4 [27] and ggplot2 4.0.3.

## 3. Results

### 3.1. Participants and pain assessments

Of the 226 participants enrolled in TRACK-SCI at UCSF/ZSFG, 16 withdrew and 70 did not complete a DN4 questionnaire, leaving 140 participants with at least one DN4 assessment (Fig. 1). They contributed 596 pain-area assessments from 330 interviews. One pain area was reported at 140 interviews, two at 114, and three at 76. The pain assessment was offered at every interview, and because the DN4 characterizes a reported pain, only 6 of the 596 assessments recorded no current pain. Forty-two participants were assessed only in hospital, 56 only at follow-up, and 42 in both phases. During the hospital stay, 112 pain-area assessments from 51 participants fell within 0 to 3 days of injury and 135 from 57 participants within 4 to 14 days, and at follow-up 133, 117, and 99 assessments were obtained from 74, 68, and 56 participants at 3, 6, and 12 months. Cohort characteristics are summarized for all participants and by assessment phase in Table 1. The cohort was predominantly cervical (77.9%) and incomplete, with AIS D the most common grade at the last in-hospital examination (60.7%), and the grade could not be determined for 11 participants. The groups contributing to each phase were similar in age and gender, whereas AIS D was more frequent among participants assessed in both phases (81.0%) than among those assessed only in hospital or only at follow-up (50.0% and 53.6%), consistent with the greater participation of less severely injured patients in TRACK-SCI follow-up.

**Figure 1.**
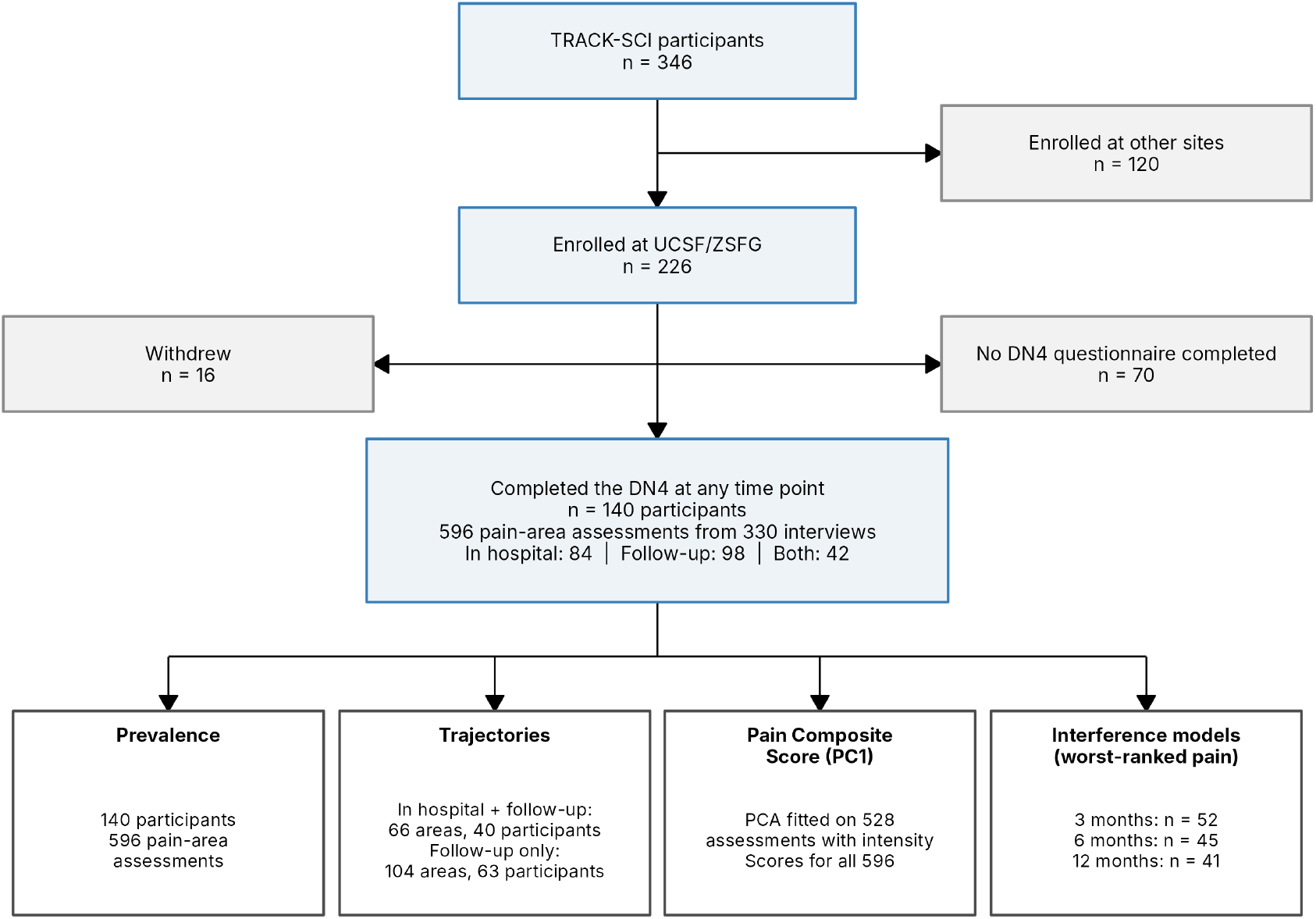
Participant flow. Of the 346 participants enrolled in TRACK-SCI, 226 were enrolled at UCSF/ZSFG and 120 at other sites. At UCSF/ZSFG, 16 participants withdrew and 70 did not complete a DN4 questionnaire, and 140 completed the DN4 at one or more time points. These participants contributed 596 pain-area assessments from 330 interviews, with in-hospital assessments for 84 participants, follow-up assessments for 98, and both for 42. The lower row gives the samples used in each analysis. Prevalence was derived from all 140 participants and 596 assessments, and the Pain Composite Score from the 528 assessments with a recorded intensity, with scores computed for all 596. Trajectories were classified for 66 pain areas from 40 participants with in-hospital and follow-up assessments and for 104 pain areas from 63 participants with at least two follow-up assessments. The interference models used the worst-ranked pain of 52, 45, and 41 participants at 3, 6, and 12 months.

### 3.2. Prevalence of DN4i-NP from the hyperacute phase

To examine whether neuropathic characteristics are already present early after injury, contrary to the view that neuropathic pain develops over months, we determined the proportion of pain areas and of participants with DN4i-NP at each time point (Fig. 2A). Within 0 to 3 days of injury, 48 of 98 pain areas (49.0%, 95% CI 40.6 to 57.4) met the DN4i threshold, and 38 of 51 participants (74.5%, 60.4 to 85.7) had at least one pain area that did. The proportion of pain areas with DN4i-NP changed little over the following year, ranging from 46.6% to 59.1% at the later time points and reaching 52.5% (40.4 to 63.8) at 12 months. The proportion of participants with at least one DN4i-NP pain area declined modestly, to 60.7% (46.8 to 73.5) at 12 months. Participant-level prevalence exceeded pain-area prevalence at every time point because participants often reported pains of different character, and at 109 of the 190 interviews with more than one pain area (57.4%) at least one pain area met the threshold while another did not (Fig. 2B). More than a quarter of all assessments (26.7%) scored 2 or 3, within one item of the threshold. DN4i-NP was therefore present from the hyperacute phase, and a considerable share of assessments lay close enough to the threshold for a single item to change their classification.

**Figure 2.**
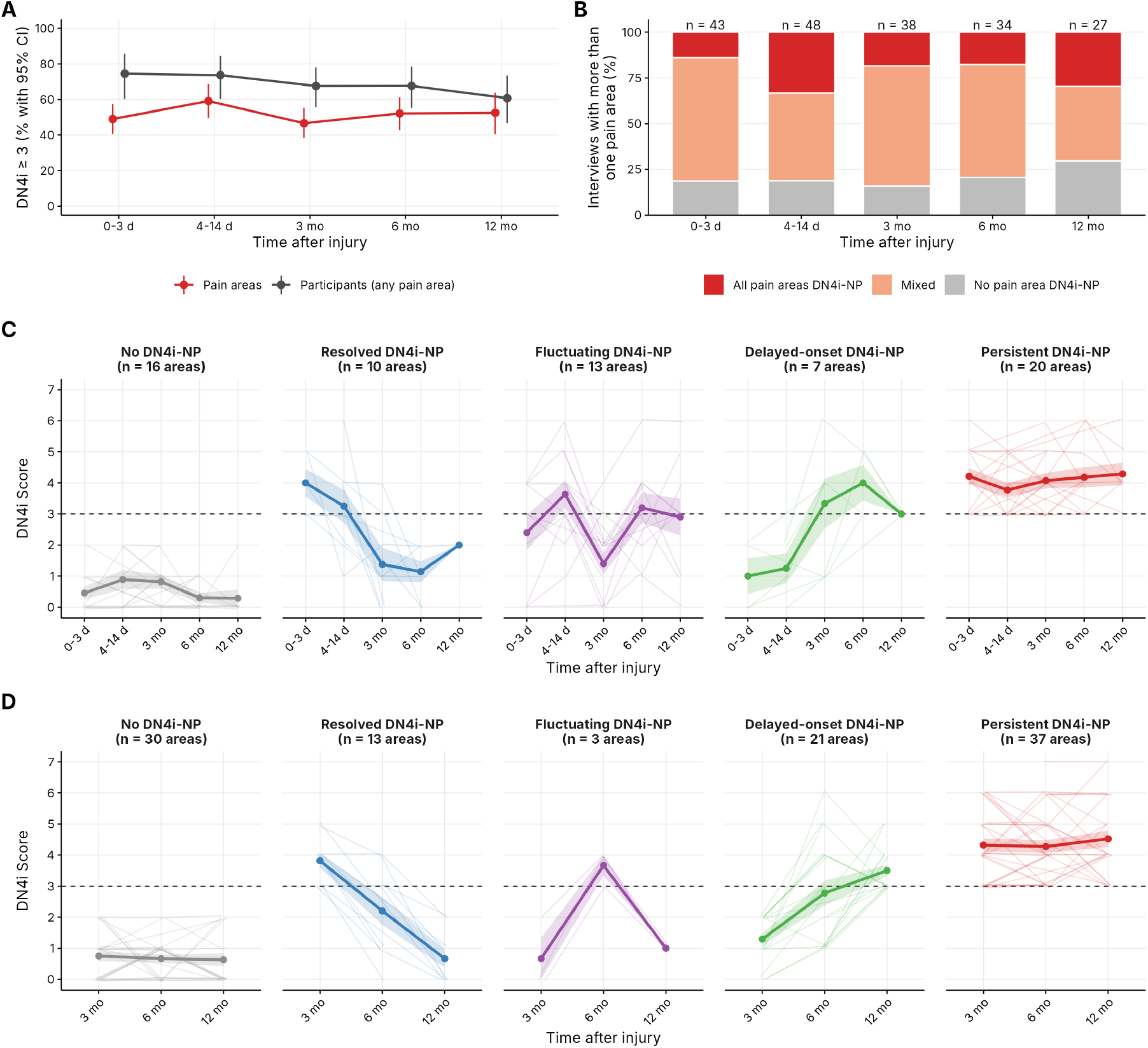
Prevalence and trajectories of DN4i-NP in the first year after SCI. (A) Percentage of pain areas and participants with a DN4 interview score of 3 or more (DN4i-NP) at 0 to 3 days, 4 to 14 days, 3 months, 6 months, and 12 months after injury. Each pain area was counted once per time bin and considered positive when any of its assessments in that bin was positive (red, n = 98, 115, 133, 117, and 99), and participants were classified as positive when any of their pain areas was positive (black, n = 51, 57, 74, 68, and 56). Points show percentages, and bars show 95% confidence intervals from a bootstrap that resampled participants for pain areas and exact binomial intervals for participants. Points are offset horizontally for legibility. (B) Interviews with more than one pain area at each time point, classified as having all pain areas DN4i-NP (red), a mix of DN4i-NP and non-NP pain areas (light red), or no DN4i-NP pain area (gray), with the number of interviews above each bar. (C, D) DN4i trajectories, with the pain area as the unit of analysis. The status sequence of each area consisted of its status in each time bin in order, with a bin considered positive when any assessment within it was positive. Areas with no change in status were classified as No DN4i-NP (below the threshold throughout) or Persistent DN4i-NP (at or above the threshold throughout), areas with a single change as Resolved DN4i-NP (positive to negative) or Delayed-onset DN4i-NP (negative to positive), and areas with two or more changes as Fluctuating DN4i-NP. Faint lines show individual pain areas (slightly jittered) and join the highest DN4i score of each area in each time bin, the value that determines the status of the bin. Thick lines and points show the mean of these values at each time point, and shaded bands show ±1 standard error. The dashed line marks the DN4i threshold of 3, and panel titles give the number of pain areas in each trajectory. Panel C includes pain areas with an in-hospital assessment (0 to 14 days) and at least one follow-up assessment (66 areas from 40 participants). Panel D includes pain areas with at least two of the three follow-up assessments at 3, 6, and 12 months, regardless of in-hospital assessment (104 areas from 63 participants).

### 3.3. Trajectories of individual pain areas

To examine whether DN4i-NP persists once present, we followed each pain area from the hospital stay through the first year. Sixty-six pain areas from 40 participants had both inhospital and follow-up assessments (Table 2, Fig. 2C), and the assessments of each participant are shown in Fig. S1. Persistent DN4i-NP was the most frequent trajectory (30.3% of pain areas), followed by No DN4i-NP (24.2%), Fluctuating DN4i-NP (19.7%), Resolved DN4i-NP (15.2%) and Delayed-onset DN4i-NP (10.6%). Resolved and fluctuating courses together accounted for one third of pain areas (34.8%). The distribution was broadly similar in a sensitivity analysis in which the status of each in-hospital time point was taken from its last assessment rather than from any assessment. Because a pain area assessed at more time points has more opportunities to change, and therefore to be classified as fluctuating, we also examined each step from one assessed time point to the next. A pain area that met the threshold at one time point fell below it at the next in 30% of cases (23 of 77), and a pain area below the threshold met it at the next time point in 31% (20 of 65). Trajectories also differed between the pains of the same person. Of the 22 participants with more than one pain area, 20 had pain areas that followed different trajectories. Most pain areas came from participants with cervical injuries (55 of 66) and AIS grade D (55 of 66), which precluded a comparison of trajectories across injury characteristics.

**Table 2.** DN4i-NP trajectories by pain area.

| Trajectory | In-hospital and follow-up |  | AIS grade, pain areas, n |  |  |  |  |  | Level of injury, pain areas, n |  |  |  | Sensitivity | Follow-up only |  |
| --- | --- | --- | --- | --- | --- | --- | --- | --- | --- | --- | --- | --- | --- | --- | --- |
|  | Pain areas, n (%) | Participants, n | A | B | C | D | E | Unknown | Cervical | Thoracic | Lumbosacral | Unknown | Pain areas, n | Pain areas, n (%) | Participants, n |
| Persistent DN4i-NP | 20 (30.3) | 19 | 2 | 0 | 0 | 18 | 0 | 0 | 16 | 2 | 2 | 0 | 19 | 37 (35.6) | 30 |
| Delayed-onset DN4i-NP | 7 (10.6) | 7 | 0 | 0 | 0 | 6 | 0 | 1 | 6 | 0 | 0 | 1 | 10 | 21 (20.2) | 18 |
| Fluctuating DN4i-NP | 13 (19.7) | 11 | 0 | 0 | 2 | 11 | 0 | 0 | 12 | 0 | 1 | 0 | 11 | 3 (2.9) | 3 |
| Resolved DN4i-NP | 10 (15.2) | 10 | 1 | 1 | 1 | 7 | 0 | 0 | 8 | 1 | 1 | 0 | 9 | 13 (12.5) | 12 |
| No DN4i-NP | 16 (24.2) | 16 | 1 | 0 | 0 | 13 | 1 | 1 | 13 | 1 | 1 | 1 | 17 | 30 (28.8) | 25 |
| Total | 66 | 40 | 4 | 1 | 3 | 55 | 1 | 2 | 55 | 4 | 5 | 2 | 66 | 104 | 63 |

DN4i-NP, pain area with a DN4 interview score of 3 or more. The in-hospital and follow-up analysis includes pain areas with at least one in-hospital assessment (0 to 14 days) and at least one follow-up assessment, with the status of each time bin (0 to 3 days, 4 to 14 days, 3, 6, and 12 months) positive when any assessment within it was positive. AIS grade and level of injury are those at the last in-hospital examination and are counted per pain area, so that a participant with several pain areas contributes to more than one row. The sensitivity analysis takes the status of each in-hospital bin from its last assessment. The follow-up-only analysis includes pain areas with at least two of the 3-, 6-, and 12-month assessments. AIS, American Spinal Injury Association Impairment Scale.

Among 104 pain areas from 63 participants with at least two follow-up assessments, the same trajectories were present, with Persistent DN4i-NP again the most common (35.6%), Resolved DN4i-NP in 12.5% and Fluctuating DN4i-NP rare (2.9%) (Table 2, Fig. 2D). Twelve of the 13 Fluctuating DN4i-NP pain areas also had at least two follow-up assessments, and eight of these were classified as Delayed-onset DN4i-NP when only follow-up interviews were considered. These pain areas met the threshold in hospital, fell below it at the first follow-up interview, at 3 months in six of the eight, and met it again thereafter. Classifying participants rather than pains obscured much of this turnover. Of 87 participants assessed at two or more time points, 46 (52.9%) had at least one DN4i-NP pain area at every time point and would have been classified as having per-

### 3.4. Pain Composite Score and dimensionality of the DN4i

To obtain a threshold-free measure of both the neuropathic character and the severity of each pain, we entered the seven DN4i items and pain intensity into a PCA. PC1 explained 33.8% of the variance, more than the 14.8% (95% CI 14.1 to 15.7%) in the permuted data (adjusted p < 0.001), whereas none of the remaining seven components exceeded its permuted VAF (all adjusted p = 1.00), so only PC1 was retained as the Pain Composite Score. Every variable loaded positively (Fig. 3A, B), most strongly tingling (0.81), pins and needles (0.79), and numbness (0.68), followed by electric shocks (0.56), burning (0.50) and painful cold (0.41), and least strongly pain intensity (0.35) and itching (0.34). PC1 increased monotonically with the DN4i total (Spearman ρ = 0.98) and resolved the eight DN4i levels into 304 distinct values, with item weights and pain intensity separating pains within each DN4i level (Fig. 3C). Its association with pain intensity was weaker (ρ = 0.37), and the DN4i total and pain intensity were only modestly related (ρ = 0.23) (Fig. 3D).

**Figure 3.**
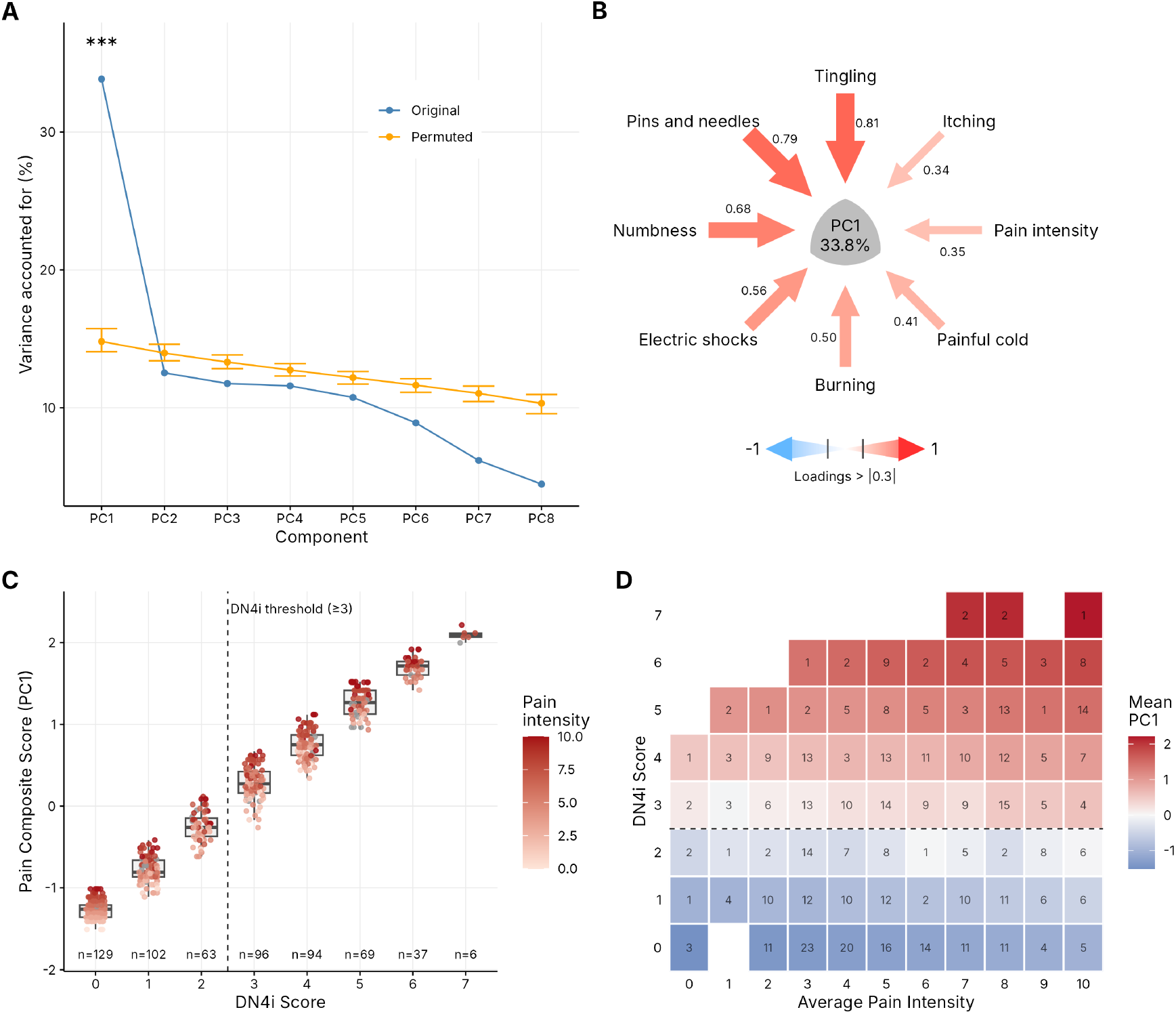
Dimensionality of the DN4i and construction of the Pain Composite Score (PC1). Principal component analysis of the seven DN4i interview items and average pain intensity in the past week (528 pain-area assessments with a recorded intensity). (A) Variance accounted for (VAF) by each of the eight components in the observed data (blue), compared with its mean and 95% confidence interval across 10,000 permutations of the data (orange, syndRomics). Only the first component exceeded the permuted distribution (PC1 33.8% versus 14.8%, Benjamini-Hochberg-adjusted p < 0.001, marked ***), whereas none of the other components did (adjusted p = 1.00). (B) Syndromic plot of PC1. Arrows point from each variable to the component, with width and color proportional to the standardized loading, whose value is printed beside each arrow, and the center gives the VAF. All eight variables loaded positively and above the plotting cutoff of 0.3, most strongly tingling (0.81), pins and needles (0.79), and numbness (0.68), while pain intensity loaded at 0.35. (C) PC1 at each DN4i score. Boxes show the median and interquartile range, with whiskers extending to 1.5 times the interquartile range. Points show individual pain-area assessments colored by average pain intensity, and gray points indicate assessments without a recorded intensity. The dashed line marks the DN4i threshold of 3, and the numbers below the boxes give the number of assessments at each score. (D) Mean PC1 (tile color) for each combination of DN4i score and average pain intensity among assessments with a recorded intensity. Numbers in the tiles give the number of assessments in each combination, and the dashed line marks the DN4i threshold of 3.

To confirm that the DN4i items themselves reflect one dimension, we repeated the analysis with the seven items alone. Only the first component again exceeded chance, explaining 37.5% of the variance versus 16.5% (95% CI 15.7 to 17.5%) in the permuted data (adjusted p < 0.001, all other components adjusted p = 1.00), and the result was the same with one pain area per participant (n = 140, 38.8% versus 19.0%, adjusted p < 0.001) (Fig. S2). The summed items showed acceptable internal consistency (KR-20 = 0.71 across the 596 pain-area assessments and 0.72 with one pain area per participant). The composite therefore rests on a single graded dimension of neuropathic characteristics, to which pain intensity contributes a smaller weight.

### 3.5. Neuropathic characteristics of the worst-ranked pain

Because pain is usually assessed and treated through the pain that the patient ranks as worst, we asked whether this pain was also the pain with the most neuropathic characteristics. At the 190 interviews in which more than one pain area was reported, the participant’s worst-ranked pain had the highest PC1 in 58.9% of interviews and the highest DN4i total in 62.6%, compared with 43.3% expected under random ranking (Fig. S3A). It was the most intense pain at 76.6% of the 184 interviews with intensity ratings for every pain area. Results were similar at follow-up alone (58.6%, 61.6% and 72.9%). When the worst-ranked pain did not have the highest PC1, the difference was substantial (median 1.03 standard deviations), and at 60 of the 190 interviews (31.6%) another pain area scored at least 0.5 standard deviations higher (Fig. S3B). Participants therefore ranked their pains more by intensity than by neuropathic character, and at almost a third of interviews a more neuropathic pain was present but not ranked first.

### 3.6. Association with pain interference

To examine whether neuropathic characteristics relate to how pain affects daily life, we modeled PC1 of the worst-ranked pain at follow-up against interference with sleep, mood, and day-to-day activities. The interference models included 136 to 138 worst-ranked pain assessments from 73 to 74 participants, and the participant random intercept accounted for 37% to 41% of the variance in PC1. Higher PC1 was associated with greater sleep interference (averaged slope 0.082 standard deviations of PC1 per interference point, 95% CI 0.033 to 0.131, F = 11.38, p < 0.001) and mood interference (0.085, 0.029 to 0.141, F = 9.40, p = 0.003), whereas the association with interference with day-to-day activities did not reach significance (0.054, −0.009 to 0.117, F = 2.94, p = 0.089) (Table 3 and Fig. 4A, with the individual assessments in Fig. S4). Both significant associations remained after Benjamini-Hochberg adjustment (p = 0.003 and 0.004) and when any single participant was omitted (all p < 0.01), and the model diagnostics showed no relevant departures from normality or constant variance (Fig. S5). Neither any pain treatment nor AIS grade was associated with PC1 in these models (p = 0.72 to 0.99 and 0.47 to 0.69). The interference-by-time point interactions were not significant for sleep and mood (p = 0.23 and 0.075) and were nominally significant for day-to-day activities (p = 0.042). Descriptively, slopes were close to zero or negative at 3 months and positive at 6 and 12 months for all three domains (Fig. 4A).

**Table 3.** Linear mixed models of the Pain Composite Score (PC1) and pain interference.

| Interference domain | Observations (participants) | Averaged slope (95% CI) | p | p (BH) | Interaction p | Between-participant slope (p) | Within-participant slope (p) |
| --- | --- | --- | --- | --- | --- | --- | --- |
| Sleep | 136 (73) | 0.082 (0.033 to 0.131) | <0.001 | 0.003 | 0.23 | 0.107 (0.002) | 0.047 (0.20) |
| Mood | 138 (74) | 0.085 (0.029 to 0.141) | 0.003 | 0.004 | 0.075 | 0.119 (<0.001) | −0.004 (0.93) |
| Day-to-day activities | 138 (74) | 0.054 (−0.009 to 0.117) | 0.089 | 0.089 | 0.042 | 0.102 (0.016) | −0.018 (0.71) |

Slopes give the change in PC1, in standard deviations, per point of interference on the 0 to 10 scale. The averaged slope and its p-value come from the Type III F-test of interference in models that included interference, time point, their interaction, any pain treatment, and AIS grade, with a random intercept per participant. BH, Benjamini-Hochberg adjustment across the three domains. Between- and within-participant slopes come from models without the interaction in which interference was split into the participant mean and the deviation from it. AIS, American Spinal Injury Association Impairment Scale. CI, confidence interval.

**Figure 4.**
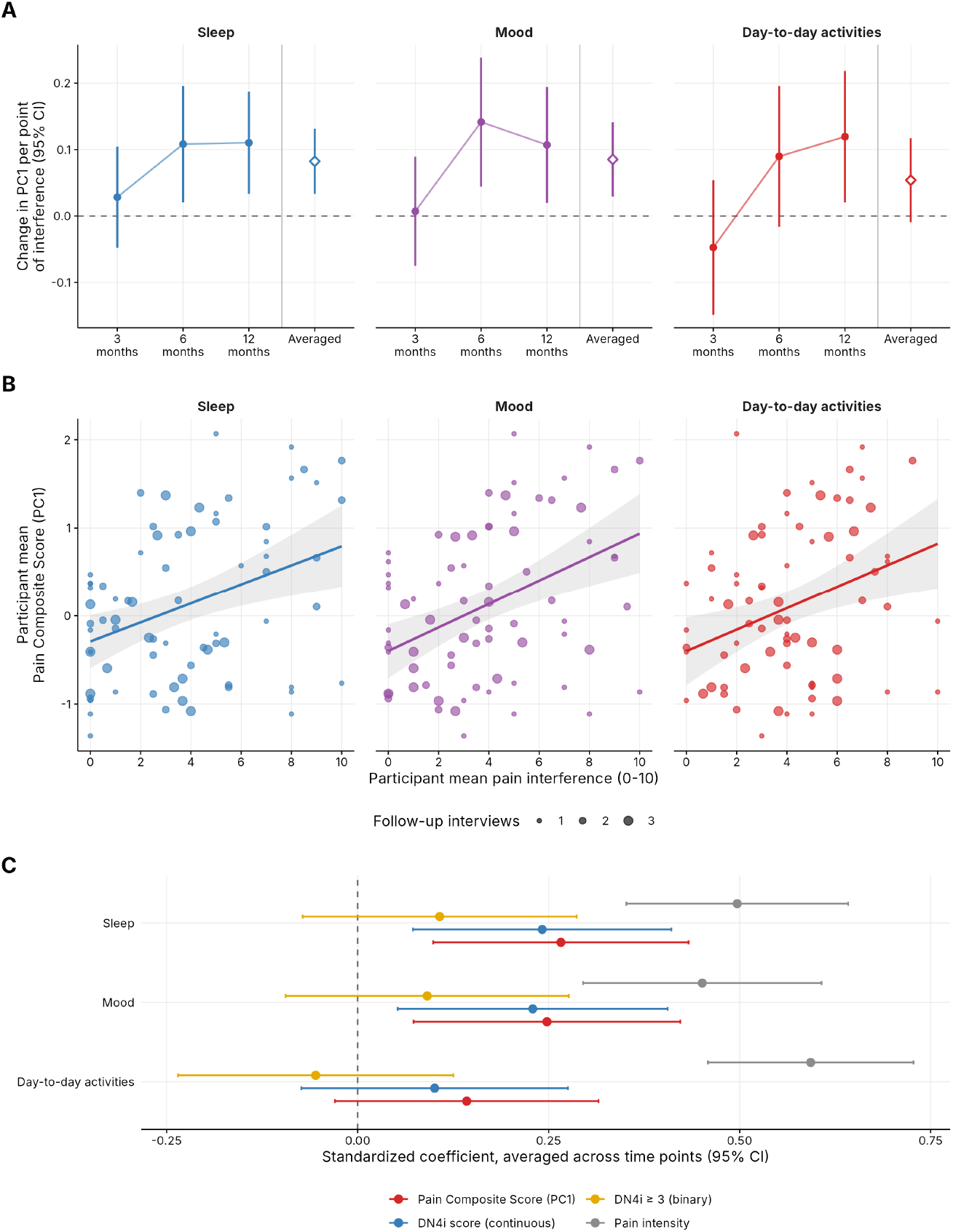
Association of the Pain Composite Score with pain interference. Linear mixed models of PC1 for the worst-ranked pain at 3, 6, and 12 months were fitted separately for interference with sleep, mood, and day-to-day activities. Each model included interference, time point, their interaction, any pain treatment, and AIS grade as fixed effects and a random intercept per participant (136 to 138 assessments from 73 to 74 participants). (A) Time point-specific slopes of PC1 on interference with 95% confidence intervals (filled points and bars), shown descriptively. To the right of the vertical separator is the slope averaged across the three time points with its 95% confidence interval (open diamond and bar), which is the quantity tested by the Type III F-test for interference. The averaged slopes were 0.082 for sleep (95% CI 0.033 to 0.131, p < 0.001), 0.085 for mood (0.029 to 0.141, p = 0.003) and 0.054 for day-to-day activities (−0.009 to 0.117, p = 0.089), with Benjamini-Hochberg-adjusted p-values of 0.003 for sleep and 0.004 for mood. The interference-by-time point interactions had p-values of 0.23, 0.075 and 0.042. (B) Mean interference and mean PC1 for each participant across follow-up interviews, with point size indicating the number of interviews contributing. The line and gray band show an interview-weighted least-squares fit and its 95% confidence interval and are included for display. When interference was decomposed into a between-participant component (the participant mean) and a within-participant component (each interview’s deviation from that mean), the association was carried by differences between participants. Between-participant coefficients were 0.107 for sleep (p = 0.002), 0.119 for mood (p < 0.001) and 0.102 for day-to-day activities (p = 0.016), and within-participant coefficients were 0.047 (p = 0.20), −0.004 (p = 0.93) and −0.018 (p = 0.71), respectively. (C) Standardized slopes averaged across time points with 95% confidence intervals. These come from the models in (A) refitted in a common sample (134 to 136 assessments), with standardized interference and outcome. The outcome was PC1 (red), the continuous DN4i score (blue), the binary DN4i classification of a score of 3 or more (yellow) or average pain intensity (gray). PC1 and the continuous DN4i score were associated with sleep interference (0.27 and 0.24, p = 0.002 and 0.005) and mood interference (0.25 and 0.23, p = 0.005 and 0.010), and neither was significantly associated with interference with day-to-day activities. The binary classification was not associated with any domain (p *≥* 0.23), whereas pain intensity was associated with all three (0.50, 0.45 and 0.59, all p < 0.001).

In the model without the interaction, the sleep and mood associations were unchanged (0.080, p = 0.001, and 0.078, p = 0.006) and the day-to-day activities association remained short of significance (0.052, p = 0.11). Decomposition of interference showed that the associations were carried by differences between participants (sleep 0.107, mood 0.119 and day-to-day activities 0.102, with p = 0.002, p < 0.001 and p = 0.016, Fig. 4B), whereas changes in a participant’s interference across interviews were not associated with changes in PC1 (sleep 0.047, mood −0.004 and day-to-day activities −0.018, with p = 0.20, 0.93 and 0.71). Participants whose pain had more neuropathic characteristics therefore reported more sleep and mood interference, whereas changes in interference within a participant were not accompanied by changes in PC1.

### 3.7. Continuous scoring, binary classification, and pain intensity

To determine whether these associations depend on how neuropathic characteristics are scored, and how they relate to those of pain intensity, we refitted the models in standardized form with PC1, the DN4i total, the binary DN4i classification, or pain intensity as the outcome. In these models, fitted to a common sample of 134 to 136 assessments, PC1 and the continuous DN4i total showed similar associations with sleep interference (standardized coefficients 0.27 and 0.24, p = 0.002 and 0.005) and mood interference (0.25 and 0.23, p = 0.005 and 0.010), and neither was significantly associated with interference with day-to-day activities (0.14 and 0.10, p = 0.099 and 0.25) (Fig. 4C). The binary DN4i classification was not associated with any domain (sleep 0.11, mood 0.09 and day-to-day activities −0.05, with p = 0.23, 0.33 and 0.54). Pain intensity showed stronger associations than PC1 for all three domains (sleep 0.50, mood 0.45 and day-to-day activities 0.59, all p < 0.001).

When interference was modeled with pain intensity and PC1 entered together, intensity remained strongly associated with all three domains (standardized coefficients 0.54, 0.45 and 0.66, all p < 0.001), and PC1 remained independently associated with sleep (0.17, p = 0.032) but not with mood (0.12, p = 0.15) or day-to-day activities (0.00, p = 0.95). The DN4i total, which does not include intensity, gave the same result when entered in place of PC1 (0.18 for sleep with p = 0.018, 0.14 for mood with p = 0.082 and 0.02 for day-to-day activities with p = 0.79). PC1 and intensity were only weakly correlated for the worstranked pain (ρ = 0.29). The number of pain areas was independently associated with mood interference (0.19, p = 0.013) but not with sleep (p = 0.38) or day-to-day activities (p = 0.56). Graded scoring thus retained associations with interference that the binary classification did not show, although pain intensity was the strongest correlate of interference and PC1 contributed independently only to sleep interference.

### 8.3. Pain location and opioid use

Because at-level and below-level pain may arise from different mechanisms and opioid use may modify pain reports, we explored PC1 by pain location and opioid use. Pain location relative to the NLI could be classified for pain area 1 at 65 of 70 follow-up interviews at 6 and 12 months (49 of the 65 were the worst-ranked pain). Thirteen pain areas straddled the lower edge of the at-level window and were classified as at-level. At 6 months, 5 pain areas were above, 19 at, and 15 below level, and at 12 months 3 were above, 12 at, and 11 below level. At 12 months, below-level pain areas tended to have higher PC1 than at-level areas (median 0.56 and 0.08), whereas at 6 months the two were similar (0.23 and 0.19). Above-level areas were too few for comparison, and the small numbers precluded formal testing (Fig. S6A).

Among participants receiving any pain treatment, confirmed absence of opioid use was recorded for only 1, 1, and 2 participants at 3, 6, and 12 months, respectively, opioid use for 16, 12, and 11 participants, and unknown medication exposure for 40, 37, and 31 participants. Pooled across time points, the median PC1 was −0.50 in the 4 participants without opioid use and 0.18 in the 29 with opioid use (Fig. S6B). These exploratory data suggest that below-level pain areas may carry more neuropathic characteristics at 12 months, whereas the medication records were too incomplete to support conclusions about opioid use.

## 4. Discussion

In this prospective SCI cohort, half of all pain areas met the DN4i threshold within three days of injury, and this proportion changed little over the first year. This stability was only apparent, however, as individual pains followed persistent, resolved, delayed-onset, and fluctuating courses, and in 20 of the 22 participants with more than one pain area, the pains of the same person followed different courses. The DN4 interview items formed a single dimension, and the threshold-free Pain Composite Score derived from them was associated with sleep and mood interference whereas the binary classification was not. At the same time, pain intensity was the strongest correlate of interference and largely determined which pain participants ranked as worst, so that the most neuropathic pain was often not the one ranked first. These findings call into question two widely held views of neuropathic pain after SCI and have implications for how neuropathic pain is defined in prognostic and biomarker studies.

Our own work on this cohort began as an effort to identify blood biomarkers that predict the development of neuropathic pain, a design that appeared straightforward under the assumption that neuropathic pain persists once it has developed. Participants could then be labeled as developing or not developing neuropathic pain, and early molecular profiles compared between the two groups. Our data show why such a label is difficult to assign after SCI. At more than half of the interviews at which participants reported several pains, one pain met the DN4i threshold while another did not, so that the participant’s classification depended on which pain was considered. Followed over time, a third of participants changed classification at least once, and assessments close to the threshold could cross it between interviews without a meaningful change in the pain itself. A model can predict only as well as its outcome is defined, and an outcome that changes with the pain chosen and the interview selected limits what any biomarker can achieve.

### 4.1. Early onset and variable course of neuropathic characteristics

The view that neuropathic pain develops over the months after SCI has been challenged by prospective work showing that it can be detected in the worst pain within days [34], is present in about 30% of patients on admission to rehabilitation [10], and is preceded by altered pain processing in the first weeks [12,19]. Outside SCI, acute neuropathic pain is recognized after surgery and trauma, where it is often overlooked although it may progress to persistent pain, and screening questionnaires have not been validated in this early phase [18]. By assessing every pain area rather than the worst pain alone, we found that half of all pain areas and three quarters of participants met the DN4i threshold within three days, even with the interview items alone. Neuropathic characteristics may therefore be more common early after injury than worst-pain assessments suggest, and the later onset reported in earlier studies may partly reflect when patients were assessed rather than when their pain changed.

The stable prevalence among pain areas over the first year and the predominance of persistent courses appear to support the second view, that neuropathic pain persists once established, as in pooled cohort data [44]. Following individual pains, however, revealed considerable turnover beneath this stability, much as reported for pain intensity [7]. Classifying the patient rather than the pain amplified the apparent stability, as more than half of the participants assessed at several time points had at least one DN4i-NP pain area at every time point, whereas only about a third of individual pains remained above the threshold throughout. One third of pain areas resolved or fluctuated. Together with the differing courses of pains in the same person, these findings suggest that the assumption of persistence, on which our earlier analysis of this cohort also relied when it combined the 6- and 12-month assessments [17], deserves to be revisited for individual pains.

The typical fluctuating course, above the threshold in hospital, below it at 3 months, and above it again thereafter, may reflect transient early symptoms, masking by nociceptive pain [34], or the limited validity of self-report in the first days after injury. Because at-level and below-level pain differ in their time course [15,37,38], some of these courses may also correspond to distinct mechanisms. The higher composite scores of below-level areas at 12 months in our data also deserve examination in larger cohorts.

Part of this turnover, and perhaps part of the views it challenges, may arise from the threshold itself. More than a quarter of all assessments lay within one item of the cutoff, where a change in a single item can register a little-changed pain as resolved or new. When the participant rather than the pain is classified, a person whose at-level pain resolves while a new below-level pain develops would be counted as unchanged, although both pains have changed. A graded description of each pain over time may give a more faithful account of its course.

### 4.2. A threshold-free score of neuropathic characteristics

The psychometric evaluation of the DN4 has focused almost exclusively on its accuracy at a threshold [4,6,30,31], and prognostic studies, including our earlier analysis of this cohort, have used it in the same way [17]. The only exploratory factor analysis we identified, performed with the full ten-item version in a mixed chronic pain population, found four factors and low internal consistency [39]. In our SCI cohort, by contrast, the interview items formed a single component and were internally consistent, possibly because the examination items, two of which formed a separate factor in that study, were not included. This unidimensionality justifies summing the items and supports, at least in this cohort, the use of the DN4i total as a graded measure.

The empirically derived Pain Composite Score was nearly identical to the unweighted DN4i total (ρ = 0.98), because all items loaded positively and pain intensity contributed little. Rather than making the composite redundant, this convergence indicates that a simple item count captures the graded information in the items, and the unweighted total offers fixed weights for prediction and biomarker models applied to new cohorts. The high loading of numbness nevertheless calls for caution, because this item may reflect sensory loss rather than neuropathic pain. Item response modeling, as has been applied to painDE-TECT [33], would establish whether the total also behaves as an interval-level measure.

Graded scoring also related more closely to daily life, as continuously scored neuropathic characteristics were associated with sleep and mood interference, whereas the binary classification of the same items was not. In SCI, where neuropathic, nociceptive, and possibly nociplastic mechanisms frequently coexist [23], a threshold that treats a pain with three neuropathic features in the same way as a pain with seven appears to discard clinically meaningful information.

### 4.3. Pain intensity and interference beyond neuropathic classification

Pain intensity was associated with interference two to four times as strongly as the neuropathic character of the pain scored with PC1, although the neuropathic association with sleep interference persisted, at roughly a third of the strength of intensity, once intensity was accounted for. Participants also ranked their pains largely by intensity, as the worst-ranked pain was the most intense pain at three quarters of interviews but the most neuropathic pain at only 59%. These findings agree with a large SCI Model Systems survey, in which neuropathic pain was more interfering across participants but, within individuals who had both pain types, differed only in sleep interference [14].

In our data, too, the associations reflected differences between participants rather than changes within them, although few participants contributed enough interviews to assess responsiveness to change. The associations were also weakest at 3 months, when interference may reflect the injury and early recovery more than the character of pain. Although intensity and interference are both self-reported and may share reporting tendencies, the results suggest that how much pain hurts matters most for daily life, that its neuropathic character makes a smaller but independent contribution to sleep, and that treatment directed at the most neuropathic pain may miss the pain that limits the patient most. The association between the number of pain areas and mood interference further suggests that the overall burden of pain, rather than any single pain, shapes how patients feel.

### 4.4. Implications for prognostic and biomarker studies

The instability of the binary label led us to a different target. Rather than asking whether a patient has neuropathic pain, a prognostic or biomarker study may ask to what extent a pain bears neuropathic features, how intense it is, and how much it limits daily life, and treat these as graded rather than binary quantities. The Pain Composite Score was derived with this aim, to combine neuropathic characteristics and intensity into a threshold-free measure that reflects the pain patients care about, and, as discussed above, it related to daily life where the binary classification did not. Graded outcomes also retain the statistical power that dichotomization discards [1], which matters in cohorts of the size typical of acute SCI research. The need for such patient-centered targets is underlined by the earlier finding that quality of life in this cohort was associated with depression and social roles rather than with injury severity [21], which shows that the classifications used to stratify patients can capture poorly what matters most to them.

Biomarker discovery in SCI pain has so far targeted mainly the presence or mechanistic phenotype of neuropathic pain [45]. Early blood transcriptomic and routine laboratory profiles already carry information on injury severity and outcome in TRACK-SCI participants [26,32], and a circulating microRNA has been associated with neuropathic pain severity modeled as a graded outcome [24], which suggests that threshold-free pain phenotypes are a feasible target for such studies. The Pain Composite Score is an initial step toward a measure of life-limiting pain, by which we mean pain that interferes with daily activities, mood, or sleep. In its current form, however, it is dominated by neuropathic characteristics and will require refinement, for example through greater weighting of intensity and the inclusion of interference, and validation in independent cohorts.

### 4.5. Limitations

This study has several limitations. Because most assessments were performed by telephone, the three DN4 examination items, hypoesthesia to touch and to pinprick and pain evoked by brushing, could not be obtained, and the DN4i has not been validated in SCI against a clinical reference standard such as the grading system of the Neuropathic Pain Special Interest Group [16], a role the patient-reported ISCIP pain type could not fill. The analysis was restricted to a single site, a third of the participants who remained in the study did not complete a DN4 questionnaire, and many contributed assessments in only one phase, so that the cohort may not represent all patients with SCI. The modest number of pain areas followed from the acute phase means that the less frequent trajectories should be interpreted with caution, and matching pain areas across interviews by their reported location may occasionally have linked different pains. In addition, pain areas were assessed at different numbers of time points, and a pain assessed more often had more chances to change and therefore to be classified as fluctuating. For this reason, we also reported how often a pain changed status from one time point to the next, a measure that does not depend on the number of assessments. Because the DN4 characterizes a reported pain, the participant-level prevalence refers to participants with pain at each time point rather than to the whole cohort. Interference was rated for pain overall rather than for each pain area, which may have weakened associations with the characteristics of a single pain, a limitation that version 3.0 of the International SCI Pain Basic Data Set now addresses [47]. Finally, medication use was recorded at every follow-up interview, but participants differed in how precisely they could report it, and opioid exposure remained uncertain at many interviews. Although opioid administration early after SCI may influence later pain in experimental models [48], too few treated participants had confirmed absence of opioid use to examine such effects.

### 4.6. Conclusions

In this cohort, neuropathic characteristics of pain after SCI were present from the first days after injury, did not invariably persist, and often differed between the pains of the same person, so that whether a patient was classified as having neuropathic pain depended on which pain and which interview were considered. A threshold-free composite of neuropathic characteristics and intensity was associated with sleep and mood interference where the binary classification was not, although pain intensity remained the strongest correlate of interference. Graded, pain-level measures that combine the character, intensity, and course of each pain may therefore offer a more stable and more informative target for prognostic and biomarker studies of life-limiting pain than a binary classification of the patient.

## Acknowledgements

The authors would like to thank all previous clinical research coordinators for their contributions during the early phases of the TRACK-SCI study, and in particular Dr. Rachel Tsolinas, Leigh Thomas, and Nicole Lai for their dedication throughout the initial years of the study. We also extend our sincere gratitude to SCI advocate Chris Barr for his continued engagement with the study and for his valuable contributions to both TRACK-SCI and this project specifically.

## Data availability

Data from this study will be deposited in the Open Data Commons for Spinal Cord Injury (odc-sci.org) under a Creative Commons Attribution license (CC-BY 4.0).

## Supplementary material

**Figure S1.**
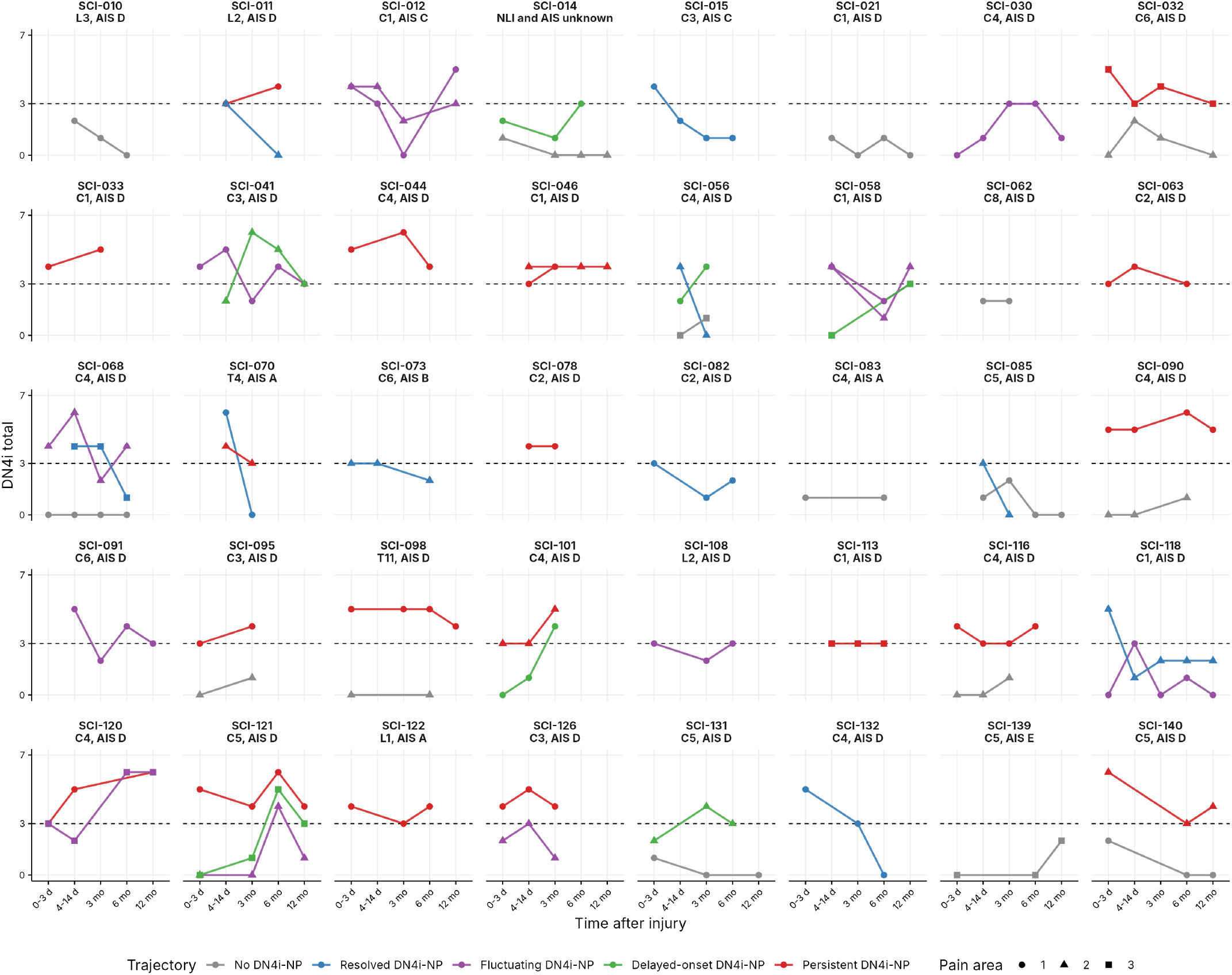
DN4i assessments of each participant in the trajectory analysis. Each panel shows one of the 40 participants with at least one pain area assessed both in hospital and at follow-up (66 pain areas), labeled with a random participant identifier and the neurological level of injury (NLI) and AIS grade at the last in-hospital examination. Each pain area is shown with one point per time bin, the highest DN4i score in that bin, which is the value that determines the status of the bin, so that a bin with several in-hospital assessments is positive when any of them is. Point shape identifies pain areas 1, 2, and 3, and lines are colored by the trajectory of the pain area as in Figure 2C. The dashed line marks the DN4i threshold of 3, and a score on the line counts as DN4i-NP.

**Figure S2.**
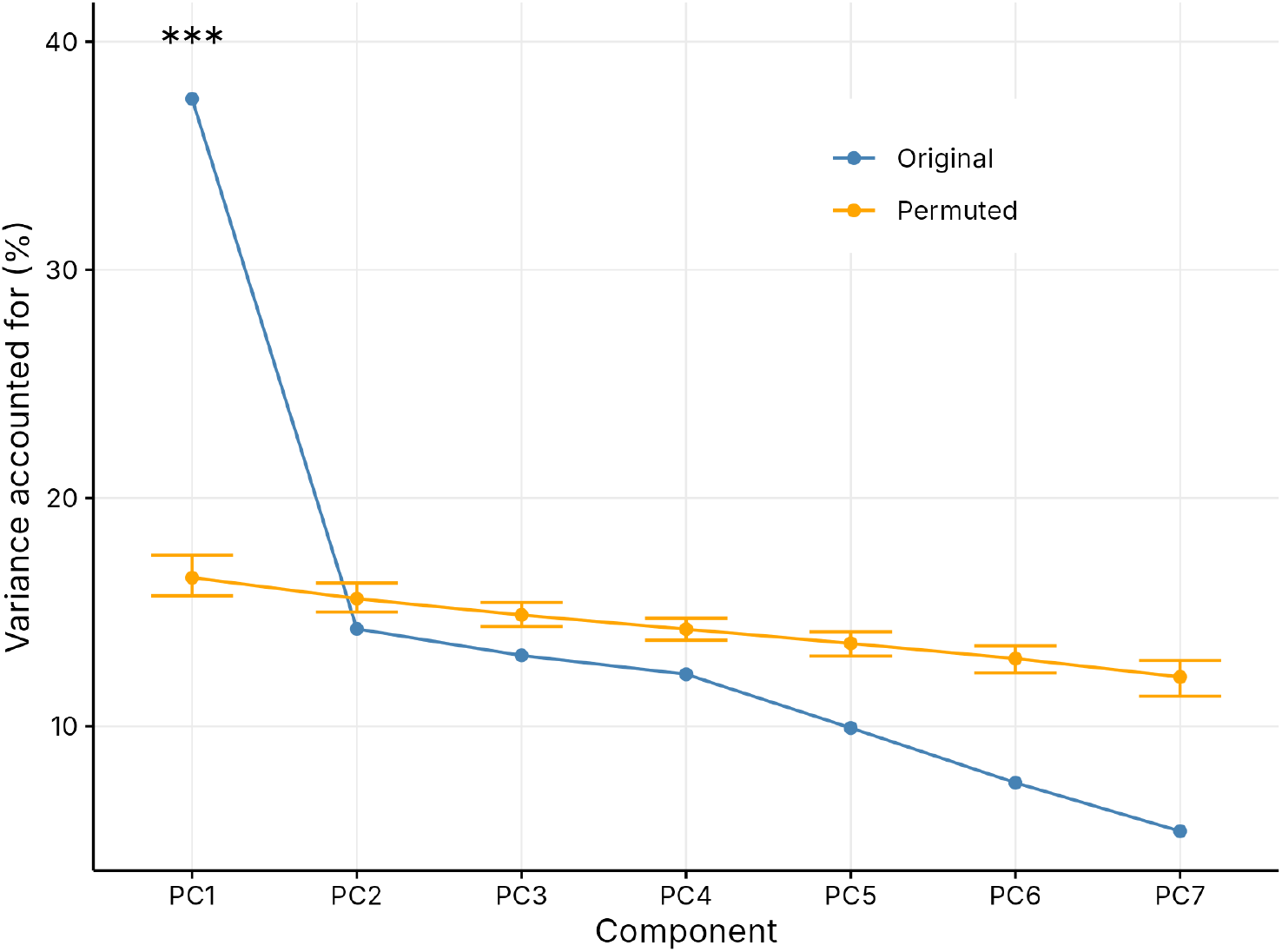
Permutation test of the principal component analysis of the seven DN4i items. The analysis included the seven DN4i interview items without pain intensity (596 pain-area assessments). The VAF by each of the seven components in the observed data (blue) is compared with its mean and 95% confidence interval across 10,000 permutations (orange, syndRomics). Only the first component exceeded the permuted distribution (PC1 37.5% versus 16.5%, adjusted p < 0.001, marked ***), whereas none of the other components did (all adjusted p = 1.00). The result was unchanged when one pain area per participant was analyzed, using the first assessment of the worst-ranked pain (n = 140, PC1 38.8% versus 19.0%, adjusted p < 0.001). Internal consistency of the summed items was acceptable (KR-20 = 0.71).

**Figure S3.**
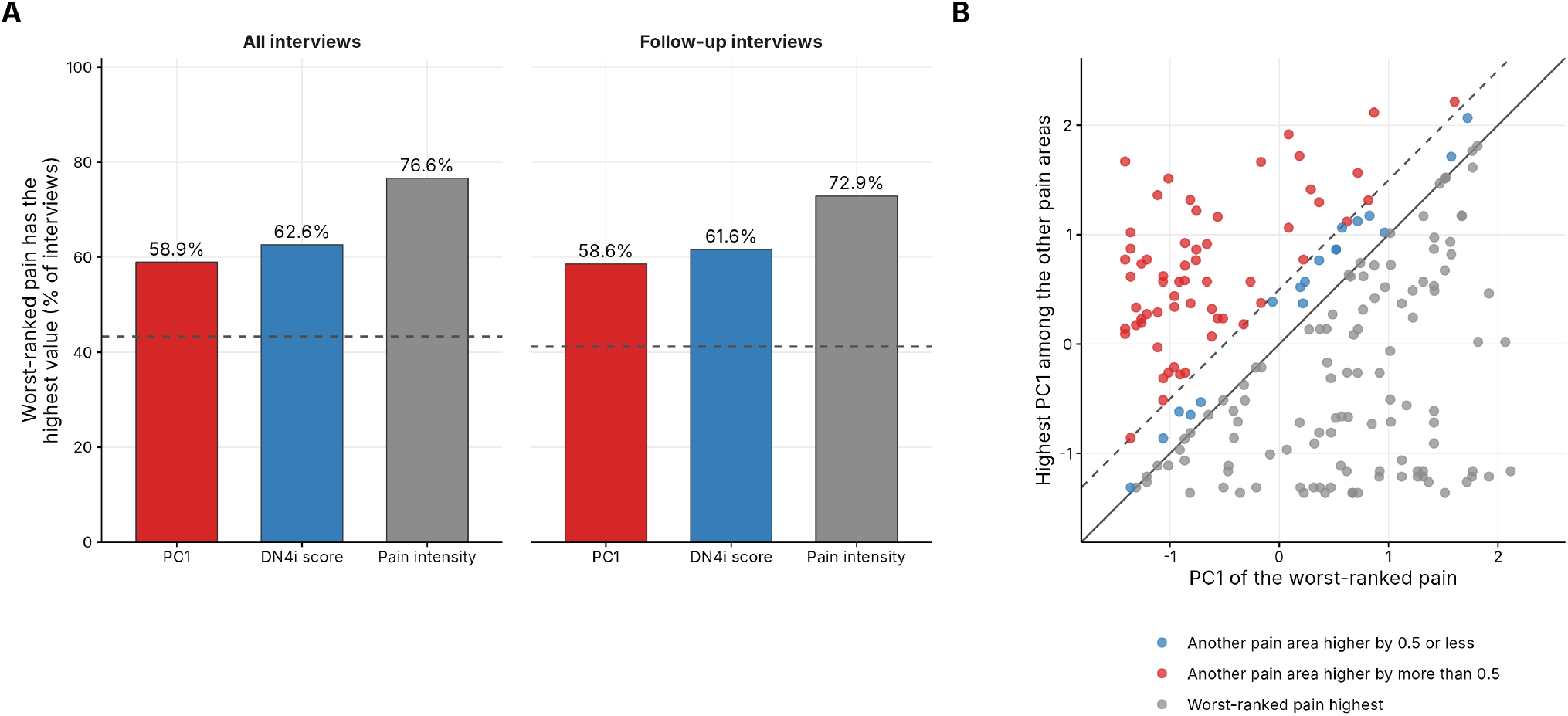
The worst-ranked pain and its neuropathic characteristics. (A) Percentage of interviews with more than one pain area at which the participant’s worst-ranked pain had the highest PC1 (red), DN4i score (blue), or average pain intensity (gray) among that participant’s pain areas, with ties counted as concordant. Results are shown for all interviews (190 for PC1 and DN4i score, 184 for intensity) and for follow-up interviews alone (99 and 96). The dashed line marks the percentage expected under random ranking, the mean of 1/k across interviews with k pain areas (43.3% for all and 41.2% for follow-up interviews). (B) PC1 of the worst-ranked pain plotted against the highest PC1 among the participant’s other pain areas at each of the 190 interviews. The solid line marks equality and the dashed line a difference of 0.5 standard deviations. Gray points mark interviews at which the worst-ranked pain had the highest PC1 (112, including ties), blue points those at which another pain area scored higher by 0.5 or less (18) and red points those at which another pain area scored more than 0.5 higher (60).

**Figure S4.**
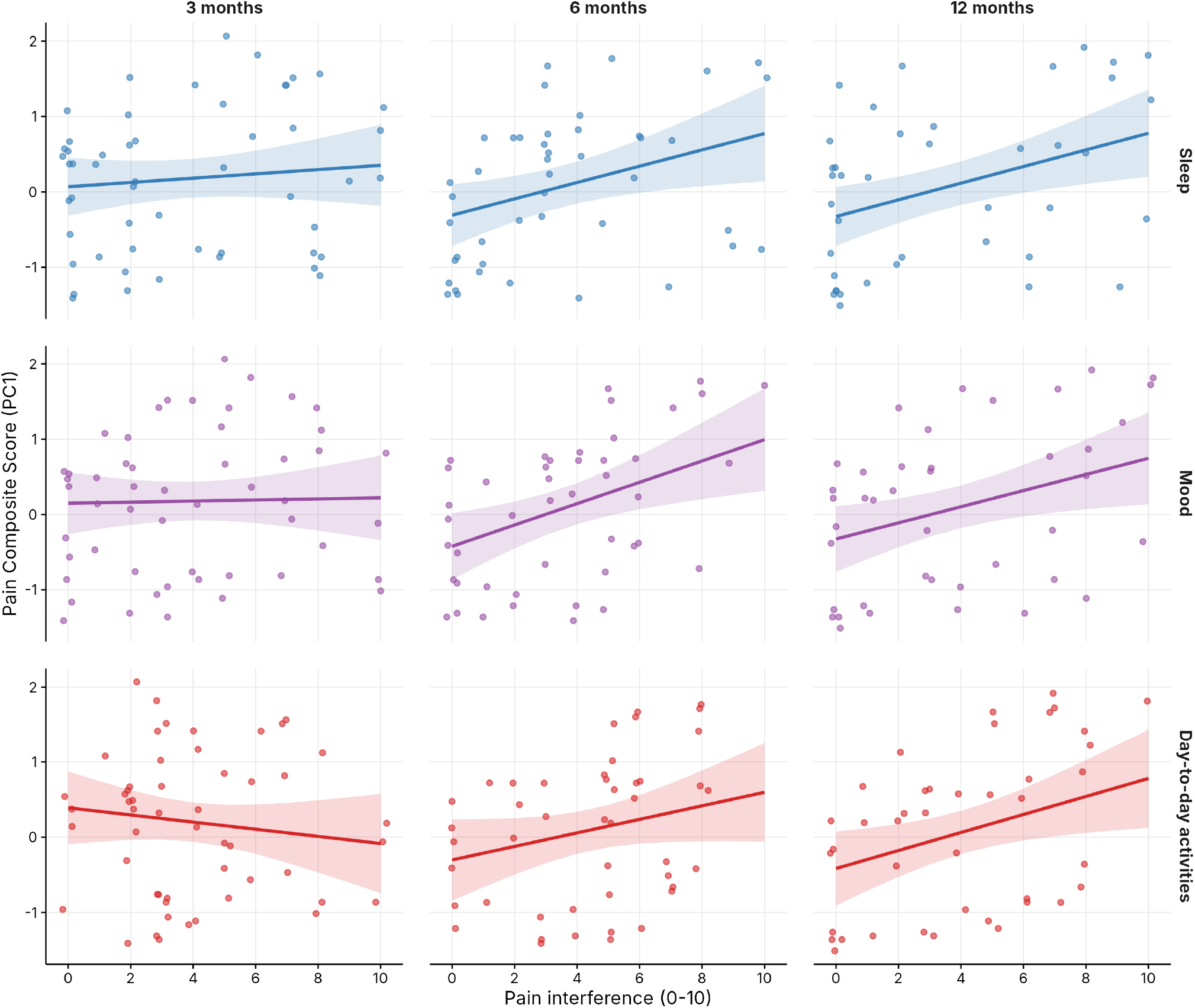
Assessment-level data underlying the interference models. PC1 of the worst-ranked pain against interference with sleep (top row), mood (middle row), and day-to-day activities (bottom row) at 3, 6, and 12 months. Points show individual assessments, jittered horizontally (n = 52, 44, and 40 for sleep and 52, 45, and 41 for mood and day-to-day activities at 3, 6, and 12 months). Lines and shaded bands show the model-predicted PC1 and its 95% confidence interval from the mixed models in Figure 4, averaged over pain treatment and AIS grade in proportion to their frequency.

**Figure S5.**
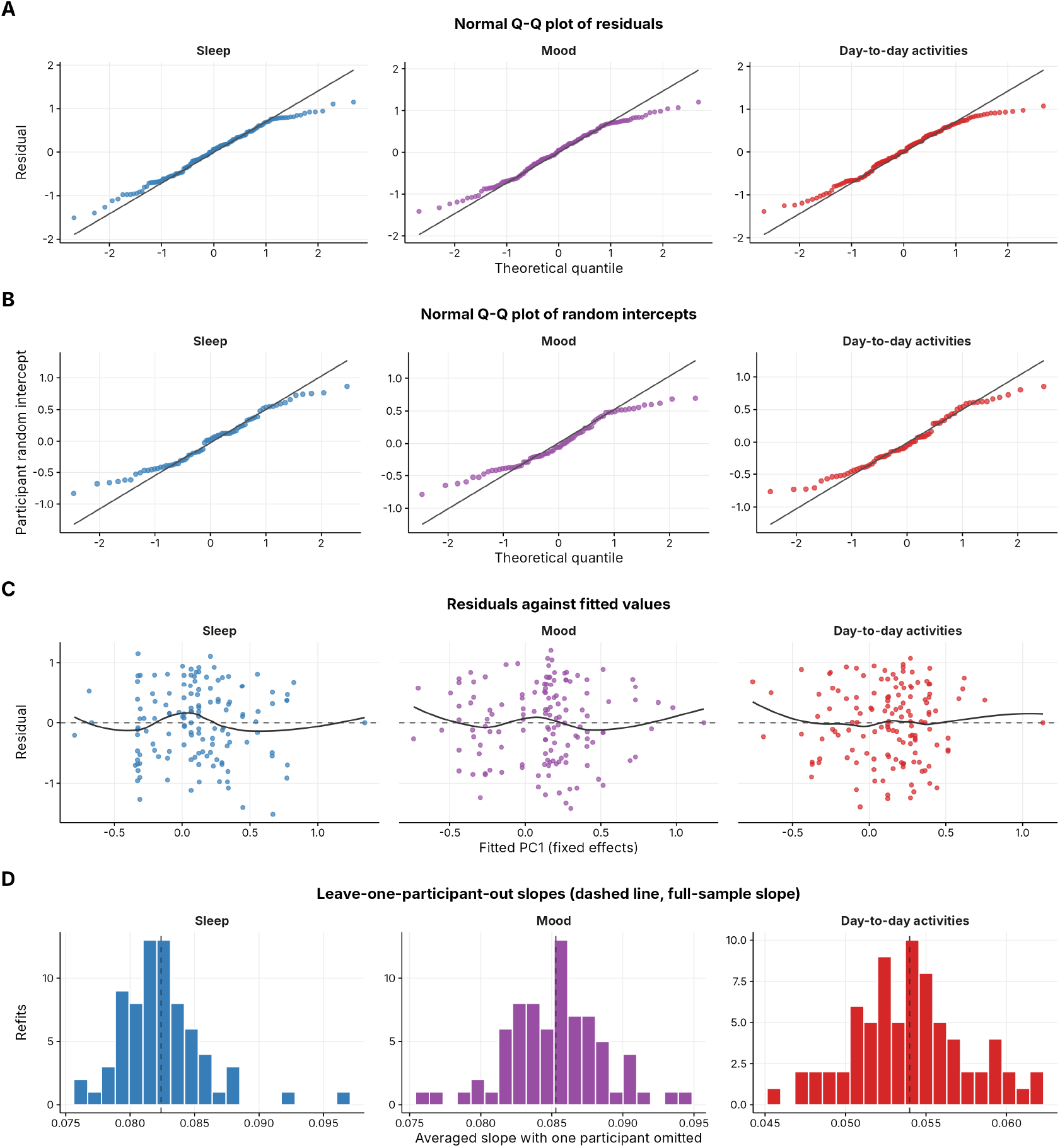
Diagnostics of the mixed models of pain interference. Each column shows the model for interference with sleep, mood, or day-to-day activities from Figure 4. (A) Normal quantile-quantile plots of the residuals. (B) Normal quantile-quantile plots of the participant random intercepts. Points close to the line indicate approximately normal distributions, and the slightly shorter tails reflect the bounded range of PC1. (C) Residuals against the PC1 values predicted from the fixed effects, with a locally weighted smoothing line, showing no trend and constant spread. (D) Averaged slope of PC1 on interference when each model was refitted with one participant omitted at a time, with the dashed line marking the slope in the full sample. The sleep and mood slopes remained significant in every refit (all p < 0.01), and the day-to-day activities slope remained non-significant (p = 0.055 to 0.16).

**Figure S6.**
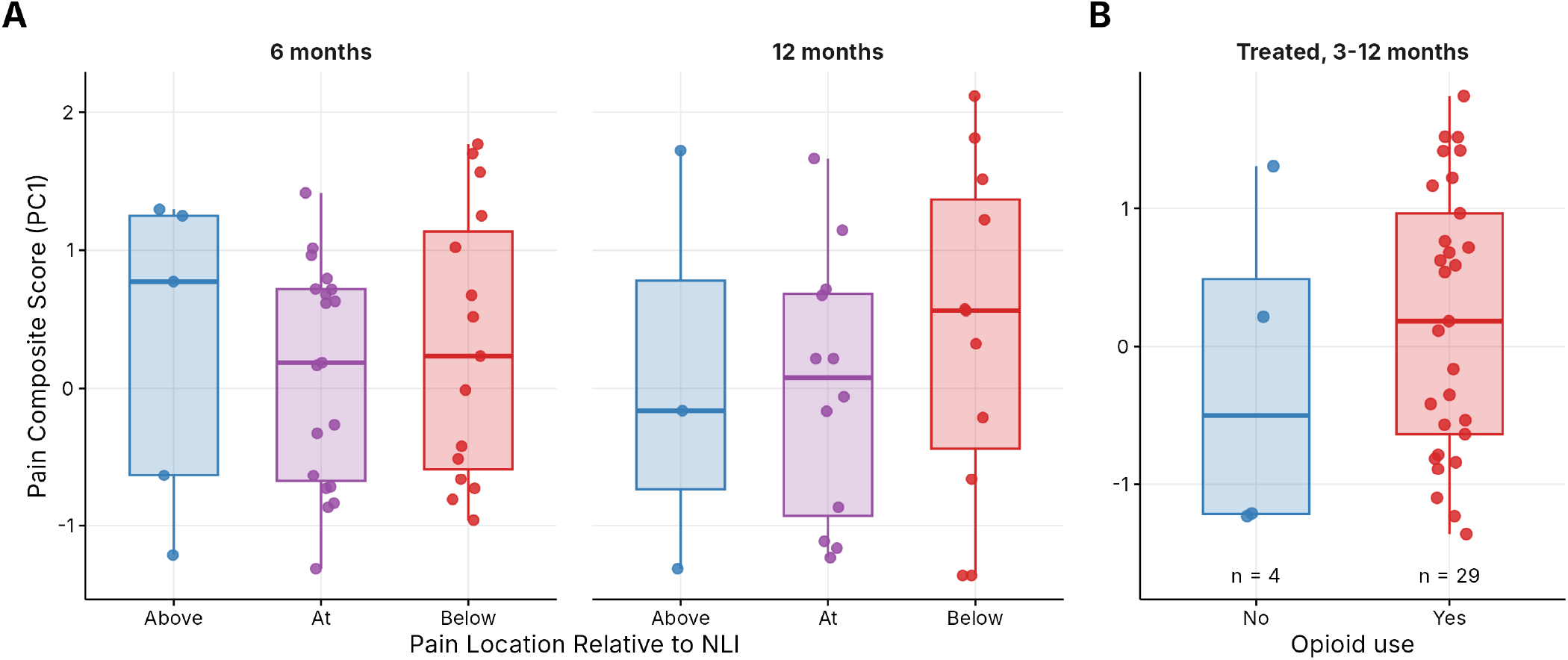
Exploratory analyses of pain location and opioid use. (A) PC1 by location of pain area 1 relative to the neurological level of injury (NLI) at 6 and 12 months. Location was classified by applying the International Spinal Cord Injury Pain (ISCIP) definition to the body regions on the pain location checklist, with the NLI from the ISNCSCI examination at the same time point as the reference level. Pain was classified as below-level when any region lay more than three dermatomes below the NLI, at-level when its regions fell between the NLI and three dermatomes below it, and above-level when all regions lay above the NLI, and pain areas with a region straddling the lower edge of the at-level window were classified as at-level (5 above, 19 at, and 15 below at 6 months, and 3 above, 12 at, and 11 below at 12 months). (B) PC1 by opioid use among participants receiving pain treatment, pooled across 3 to 12 months. Opioid use was classified at the participant level for each follow-up interview, as present when it was recorded for any pain area and as absent only when it was recorded as absent. Interviews with unknown medication exposure were excluded (40, 37, and 31 participants at 3, 6, and 12 months), and PC1 was averaged across each participant’s interviews within each category (4 participants without and 29 with confirmed opioid use, with median PC1 of −0.50 and 0.18). Boxes show the median and interquartile range, and points show individual pain areas (A) or participants (B). Given the small numbers, both analyses are presented descriptively.

